# Exposures associated with sporadic *Cryptosporidium* infection in industrialised countries: a systematic review

**DOI:** 10.1101/2022.03.08.22272080

**Authors:** Caoimhe McKerr, Wendi Shepherd, Rachel M Chalmers, Roberto Vivancos, Sarah J O’Brien, Alison Waldram, Kevin G Pollock, Robert M Christley

## Abstract

Transmission of Cryptosporidium can occur via contaminated food or water, contact with animals or other people. Exposures are often identified from outbreak investigations, but sources for sporadic disease and pathways to infection are still unclear. The aim of this review is to consolidate the literature to describe exposures associated with human cryptosporidiosis in industrialised countries.

Methods followed the recommendations made in the Cochrane Handbook for Systematic Reviews of Interventions. Three steps were used to identify the literature including electronic database searching using PubMed, Scopus and Web Of Science; reference list trawling; and an exploration of the grey literature. Quality was assessed using the Newcastle-Ottawa Scale. Screening of results was undertaken by two reviewers and data extracted using a standardised form. A narrative summary presented. Papers were included if they reported on sporadic cases and were published between 2008 and 2018. Exposures were grouped into pathways.

After full-text screening, eight articles (comprising 11 studies) were included, and seven (comprising 10 studies) were suitable for further synthesis. None of the identified grey literature was included. Four studies described case-control methods, two were case-case studies and one cross-sectional. Study year ranged from 1999 to 2017 and the studies were conducted in five, large countries in three continents: Europe (UK and the Netherlands), North America (USA, Canada), and Australia.

Included papers investigated water and animal exposures most frequently. Recreational water was not a major source of sporadic illness in this review. The person-to-person pathway represented the most consistent finding, with all three studies reporting on contact with a symptomatic individual demonstrating correlations between exposure and disease. This applied particularly to the home environment, which is increasingly understood to be a significant setting for spread of Cryptosporidium infection. Further work on this would help support public health messaging on preventing spread of disease at home.

Systematic review registration: PROSPERO number CRD42017056589.

## Introduction

*Cryptosporidium* is a protozoan parasite which can infect humans and other animals. The most prevalent species identified in humans are *Cryptosporidium parvum* and *Cryptosporidium hominis* (1,2). *Cryptosporidium* is distributed world-wide and is suspected to be one of the greatest contributors to human infectious diarrhoeal illness (3). *Cryptosporidium* infection is reported in 1-3 percent of immunocompetent patients with diarrhoea in industrialised countries and 7-20 percent in developing countries (4–8). The dissimilarities are believed to be driven by variation in exposure due to sanitation, availability of clean drinking water, infrastructure, housing, and health factors such as acquired immunity and nutrition (Putignani and Menichella, 2010). The highest prevalence is observed among children under five years old, in particular the under twos (3,10). The parasite has a complex life cycle, and characteristics such as structurally tough, resistant oocysts help favour the faecal-oral transmission route. Outbreaks via animal-to-person (*C. parvum*) contact are commonly reported, as well as those associated with indirect transmission routes through ingestion of water and food, and, less often, person-to-person (*C. hominis and C. parvum*) routes (11).

Reported exposures for both *C. parvum* and *C. hominis* often overlap, and include consumption of contaminated drinking water and exposure to recreational waters, (12–14) and food-related outbreaks (likely contaminated via water or by food handlers) (15–18). *C. parvum* is frequently associated with exposure to farm animals (19,20) and *C. hominis*, more anthroponotic, with person-to-person spread (21–24), and increased risks during foreign travel (25). However, the risk factors and associated exposures are often hypothesised/identified from outbreak investigations, but outbreaks may only represent a small proportion of cases - in the UK, less than 10% of all cases reported to national surveillance are linked to an identified outbreak (26). Additionally, surveillance systems may not always capture smaller yet significant outbreaks, and under-ascertainment of true case numbers is common (27–30). As a consequence, pathways may be under-reported and we cannot be certain that transmission routes for sporadic disease are the same as those which drive outbreaks (31). Despite case control studies which have investigated differences in risk for endemic and outbreak disease (32,33), sources for sporadic disease and pathways to infection are still unclear and a proportion of reported cases remain unexplained.

Given the absence of any systematic synthesis of reported evidence in the UK, and the low numbers of reviews in other industrialised countries, the aim of this work was to search the literature to describe exposures associated with sporadic infection with *Cryptosporidium* in industrialised countries, and to explore differences that may exist in these associations between sporadic and outbreak-related disease.

## Methods

**The full methods and approach have been previously reported** (34)

Where relevant, methods followed recommendations made in the Cochrane Handbook for Systematic Reviews of Interventions (35) and reporting followed guidance from “Preferred Reporting Items for Systematic Reviews and Meta-Analyses” (PRISMA) (36,37).

### Eligibility and inclusion

Databases were initially searched with no restriction on year of publication but analysis was later restricted to 2008-2018. There were no restrictions on language, provided the abstract was available in English for the first round of screening.

Studies conducted in industrialised countries (defined using OECD category of countries (38) additional file 1) and reporting on human subjects were included. All observational studies were included where they reported exposures and relevant quantitative results. Individual case reports were excluded as in single case descriptions do not give a picture of population epidemiology.

### Data sources

Three steps were used to identify the literature including: electronic database searching using PubMed, Scopus, and Web of Science; reference list trawling from relevant papers; and an exploration of the grey literature. The last published search date was 15th May 2018, and grey literature on 21^st^ May 2019.

### Searches

The search terms included the following MeSH terms/ keywords: (Cryptosporidium OR cryptosporidiosis) AND (epidemiolog* OR risk factors OR exposure OR transmission OR association) OR (cohort OR case-control OR “case control” OR case-crossover OR “disease outbreaks” OR meta-analysis OR longitudinal OR ecological). (Additional file 2)

### Study selection process

Results were managed using the Covidence tool (39). Screening of results and title screening was undertaken in duplicate by two reviewers, to ensure consistency in the application of the inclusion and exclusion criteria. There was a change in second reviewer during the abstract review (AW, replaced by WS) – however, the first (CMcK) and third (KP) reviewers did not change which maintained consistency. Discrepancies were discussed and re-examined, and a third reviewer was available for irreconcilable opinions on inclusion. One paper was sent to the third reviewer for decision.

### Data management

Data were extracted in duplicate using a standardised form developed in MS Access. Minimum data fields are available in Additional file 3. Data items were merged, and discrepancies discussed, prior to the agreement of which data were appropriate for analysis.

### Quality assessment

Papers were scored using the Newcastle-Ottawa Scale (NOS) (40). This instrument is well piloted and is specific to non-randomised study types. The NOS was completed independently by both reviewers for all studies. Results were then amalgamated, and areas of discrepancy discussed prior to agreement on final scores. This instrument provides an overall judgement on quality using a scoring system by evaluating three parameters (selection, comparability, and outcome) across specific domains. The maximum score for each study is eight: four stars for selection, one star for comparability and three stars for exposure/outcome domains. The studies were considered of high quality if the NOS score was 7+ stars (41), moderate quality for a score of 5 or 6 stars, and studies having fewer than five points considered at high risk of bias/poor quality (42).

### Data synthesis

Data were summarised presenting the papers’ main findings including population under study, outcome(s) measured/case definition, effect measures and reported statistics, and exposures. Exposures, depending on how they were measured, were grouped into transmission pathways, following discussion and consensus with authors. Broadly the pathways considered included water, animals, food, and person-to-person spread. We further sub-categorised exposures following discussions with the study group, in order to better represent specific known transmission risks for *Cryptosporidium*, with possible differences between species highlighting the anthroponotic versus mostly zoonotic pathways.

We proceeded with a visual and narrative analysis of results for each exposure in the transmission pathways. Where appropriate, available data were pooled in a visual analysis combining the exposures across studies using GraphPad Prism 8.3.0 (43). Where a usable effect measure was not reported in the study, but data were available, odds ratios or relative risks were calculated. Where available, the multivariable-level effect measure was used in preference to the crude to ensure where possible we were using adjusted variables which remained significant predictors of infection after controlling for covariates. All results, whether statistically significant or not, and those either showing the exposure as increasing or reducing risk of outcome, were considered in the analysis. For those studies where we used multivariable analysis, all results will have remained in the final, most parsimonious model.

### Additional exclusion criteria

Following full text review, additional restrictions on type of cases and time period were applied to refine the selection of papers taken forward for data extraction.

It became apparent to the authors that many of the included papers described (mostly) outbreaks in great detail, with very different, but specific, settings and widely variable metrics for exposures. This made it difficult to meaningfully synthesise the papers’ outcomes within the scope of this review. Additionally, outbreak-focused papers were often specific to one event, exposure, or setting, and posed the additional problem of more granular exposures and led to difficulties with quantifying direct exposure. As the main focus of the review was centred on sporadic, community cases, we applied additional criteria after full-text screening of papers to remove those describing outbreak investigations.

Additionally, the older studies were less relevant for describing contemporary risks, and often traversed changes in policy and practice (such as changes to municipal water supply regulations) which may contribute to changes in reported exposures for disease over time. Therefore, in order to focus the synthesis on the most relevant and contemporaneous data we restricted inclusion at the data extraction stage to reports within the last ten years (2008-18).

## Results

The combined initial database search retrieved 2,115 articles. An additional article was found by screening reference lists, and one pre-print was included following a routine refresh of the search terms on PubMed. This was reduced to 1,797 after removal of duplicates.

Based on title and abstract screening, 338 full-text articles were procured and retained for potential inclusion (**Error! Reference source not found**.).

After full-text screening, 208 peer-reviewed papers were excluded and the remaining 130 taken forward to data extraction. We excluded papers which were part of wider studies and which reported on the same aspects of the study to prevent inappropriate weighting to one study reported multiple times.

The application of additional selection criteria (as previously described) at this stage were two-fold:

1. To restrict the inclusion of papers to the past 10 years (2008-2018) , and
2. To restrict the review to studies of sporadic disease

These further restrictions resulted in the inclusion of eight articles (comprising 11 studies), of which seven (comprising 10 studies) were suitable for further synthesis. The single paper excluded from any quantitative analysis did not have enough information for further analysis or calculations, and no reply was received from authors to our request for additional data.

None of the identified grey literature articles were included (searched between May and Sept 2019) as they all described outbreaks and/or were outside the time period. Six relevant theses were captured but were out of area, or outside the time scope, or were part of published papers we had already captured.

### General characteristics of included studies

A total of eight papers describing 11 individual studies were included for review and potential analysis. The Fournet *et al* paper described three studies from the UK, Germany and the Netherlands: we excluded the Germany study due to lack of available data and report this paper as two separate studies from the UK and the Netherlands. Study numbers for the smaller case-case study in the Fournet (2) UK study were not reported.

The NicLochlainn *et al* paper includes only results for the overall study period, not individual years, to avoid duplicate counting.

The Tollestrup *et al* paper described one large cohort study of 600 participants, but analysed data by residence of participants across three sites: we report results for the Tollestrup paper as three individual studies.

### Risk of bias assessment and quality of the included studies

shows the NOS scores allocated to each of the articles and includes stars allocated for each of the individual domains, and reports on general strengths and weakness of the study designs. For the Tollestrup cohort studies across three sites, (Tollestrup *et al*., 2014) the overall article score was taken, as the study designs were the same and thus of identical quality.

Almost all studies (n=7) were at least of moderate quality, although some specific biases were observed in the individual domains. Non-response rate in particular, and the description of and differences in responses in participants, was of some concern in almost all of the studies. A key quality aspect was risk of bias in the ascertainment of the exposure and its comparability to case exposure. In a couple of the articles it was felt that case representativeness introduced a likely bias.

**Error! Reference source not found**. shows the main characteristics of the included studies.

Four of the studies described case-control methods, two were case-case studies and one cross-sectional. Four studies (across 2 papers) reported outcomes based on serology and thus were classed as serological studies. All but one of the studies had over 100 participants, with three comprising several thousand participants – these tended to be those that selected cases from large, nationwide surveillance databases.

Study year ranged from 1999 to 2017 and the studies were conducted in five, large countries in three continents: Europe (UK and the Netherlands), North America (USA, Canada), and Australia. The European focus on the UK and the Netherlands might be explained by differences in the approach to detection and legislation of reporting for *Cryptosporidium* and this awareness within European countries, and not necessarily reflect a true lack of cases (9).

### Transmission pathways and exposures investigated

In discussion with the review team, recognising the faecal-oral transmission route for *Cryptosporidium* infection in humans, the following transmission pathways were established capturing the predominant exposures measured in the included studies:

- **Animals** – pets and farm animal exposures.
- **Food** – eating spoiled foods, raw or fresh produce, unpasteurised beverages, and proxy exposures such as eating outside the home, killing/preparing own meat.
- **Outdoor activities/environmental exposures** – activities such as gardening and hiking/camping, and environments such as living on a farm.
- **Person-to-person spread** – contact with a case (close contact, caring/toileting capacity, and sexual contact) and general social contact/activities.
- **Water** – drinking water and recreational water contact/water sports
- **Travel** - any travel away from usual area of domestic residence.
- **Season –** season of infection, usually extracted from surveillance data.

Main findings can be found in **Error! Reference source not found**. which outlines the main exposures measured (as defined by the studies) and the main results reported with a relevant transmission pathway added. Where available, multivariable results have been used (OR=odds ratio, aOR=adjusted odds ratio).

The included papers measured over 200 exposures across the 11 studies. Exposures measured were multiple and varied in detail and definition across each of the studies, and even within studies. Generally, studies were exploratory in nature. The 2015 de Gooyer *et al* study in Australia (48) was centred on a specific hypothesis about exposure to recreational water, but the rest of the studies tended to look at myriad exposures, following the main known routes to infection for *Cryptosporidium*. Exposure to animals was a common pathway, with six studies investigating this and both livestock/farm animal exposure and pets and domestic animal contact were well investigated. Food items were investigated in five studies, and personal contact (with a case or general social contact) in seven. All bar one study investigated some water exposures, the most commonly investigated pathway for *Cryptosporidium* cases.

The Becker (2015) (51), Pintar (2009) (50), and Tollestrup (2014) (45) studies also included risk factors relating to patient/case demographics and characteristics. We include a short, narrative overview of this content.

### Clinical variables, case demographics and social factors

Pintar’s 2005-7 case-case study of the Canadian Integrated Enteric Disease Surveillance System (C-EnterNet) reported that the odds of a *Cryptosporidium* case being between six and twelve years old was five times greater than the other enteric pathogens. The non-*Cryptosporidium* controls had an older profile than the cases (mean: cases=21.7 years, controls=31.8 years, p=0.01). Among the serological studies, increased odds of seropositivity was observed with increasing age: in those over 60 years old compared to a wider baseline of under 40 years at two sites in the Tollestrup studies, and with increasing age in the Becker study (p<0.001). However, it is well known that cases exhibit a bi-modal age distribution pattern and serological studies can report increased serological positivity with increasing age (52), which may reflect amount and intensity of exposure over time, or elevated serological responses increasing likelihood of detection (53).

In the Tollestrup (b) study, the authors reported a reduced odds of positive serological response to *Cryptosporidium* antigens in people reporting higher education levels (**Error! Reference source not found**. In the higher income countries, sporadic illness is more often observed among the less deprived areas and communities (54,55). In these countries, rurality is associated with increasing wealth while more deprived areas tend to be located in the city. Additionally, associated activities such as swimming and travel are likely to be less prevalent in the more deprived areas and as such the profile of *Cryptosporidium* in relation to deprivation is converse to that in the less industrialised areas (41,56). Conversely, Becker *et al* analysed data extracted from the USA National Health and Nutritional Examination Survey (NHANES is series of surveys which examines about 5,000 persons each year in the USA and is considered to be a nationally representative sample. The NHANES interview includes demographic, socioeconomic, dietary, and health-related questions as well as medical, dental, and physiological measurements (Centers for Disease Control and Prevention (CDC), 2017) between 1999 and 2000, and found that correlates to *C. parvum* included several poverty and inequality measures including country of birth other than USA (p<0.001) and ethnic groups other than non-Hispanic whites (p<0.001). These results are supported by similar work reporting that Hispanics, African Americans, and women, all had greater odds of reporting *Cryptosporidium* seropositivity (57). Whilst relative poverty alone is unlikely to directly cause infection with *Cryptosporidium*, it might steer exposure to particular pathways or risks, or indeed reduce the availability of resources required to avoid exposure, either by personal behaviour/engagement, general health status or access to a health infrastructure (55,58). However, assessing individual risk alongside population-level characteristics often introduces fallibility, especially when using surveillance data which may introduce a bias in participants based on their access to healthcare. Further work on specific population-level characteristics would be useful and may help contextualise some of the individual-level relationships between these characteristics and exposure to infection.

Whilst it is important to note these findings, these parameters are likely to be risk factors associated with exposures to *Cryptosporidium*: we only used metrics which could be categorised into *Cryptosporidium* transmission pathways in the further analysis, thus excluding case characteristics. Three studies looked at travel as a risk for disease (any travel away from usual domestic residence) but most often it was an exclusion criterion in order to ensure investigation of endemic indigenous cases. *Cryptosporidium* is often observed as a non-viral cause of gastrointestinal illness in returning travelers (59) and travel might subject people to risk exposures. As such it is not an exposure for disease intrinsically, and susceptibility may be linked to immunity, or lack thereof, of travelers on short term holidays in endemic areas (60) as well as increased exposures in areas with lower hygiene or more frequent use of swimming pool in holiday resorts. We excluded travel from any further detailed analysis.

Additionally, there were insufficient papers reporting on season or outdoor activities in enough detail to analyse these any further

### Transmission pathways

We excluded the entire Ravel *et al* (2013) study from any further analysis as measures reported were proportion exposed rather than odds or risk, and the data were not available to enable calculation of an effect measure that was comparable with the other studies. Ravel *et al* (2013) used the C-EnterNet surveillance system to link confirmed cases of sporadic, domestically acquired *Cryptosporidium, Giardia* and amoebiasis with exposures grouped by the main pathways: Water, animal/environment, person-to-person, and exposure to high-risk food, analysing each infection separately. For cryptosporidiosis cases, travel within Canada (100%), contact with household pets (49%), and swimming (46%) were the most frequently reported exposures. The animal/environment-to-person transmission pathway remained the most important factor in *Cryptosporidium* cases’ exposure(s) (72%). This was followed by water-based transmission routes (52%) and exposure to risk foods (50%). We were unable to look at odds of *Cryptosporidium* using the other cases (amoebiasis and giardiasis) as ‘controls’, given the differences in the system characteristics and the lack of individual response data.

We further sub-categorised the transmission pathways following discussions with the study group, in order to better analyse the underlying exposures for *Cryptosporidium* (***Error! Reference source not found***.). This level of granularity in exposures helps highlight differences between anthroponotic and zoonotic pathways, important in recognising differences between infecting species and also when considering targeted public health messages. Although this made the numbers of studies within each category lower, the team felt that aggregating to top level pathways lost important detail that is imperative in understanding *Cryptosporidium*.

As no exposure included more than four studies, a traditional meta-analysis was considered inappropriate (61). However, a visual analysis allows us to easily see which studies reported on the main underlying exposures investigated in each pathway and discuss each of the results in light of the characteristics of the studies.

Ten studies (across seven papers) were included in a further analysis allocating the exposures investigated in the studies into the relevant transmission pathway (where we could allocate a main route).

All results whether statistically significant or not, and those either showing the exposure as increasing or reducing risk of outcome, were considered in the analysis. We used multivariable results where we had them for the comparable exposure. Charts show the study name, study sample size, measure of association reported, and confidence intervals. Black dots represent statistically significant results, and red insignificant. (Figures 3-12)

## Results and discussion by transmission pathway

### Transmission pathway - Animal contact

Animal contact exposure results were reported from five of the included studies. Animal exposures were categorised as livestock (n=3) and pets (n=2). **Error! Reference source not found**.

#### Farm animals

One study reported a statistically significant increased odds of disease (Pintar; OR=1.6; 95% CI=1.1-2.5) and another reported reduced odds of seropositivity (Tollestrup (a), OR=0.53; 95% CI=0.29-0.97). The remaining study reported an insignificant result below 1.0 (Fournet (1), 2013; OR=0.74, 95% CI=0.41–1.32).

The metric used in the Pintar study was “visited farm, petting zoo, or fair” which approximates animal contact but there was no detail about physical contact with animals. In their univariate analysis, they had an environmental exposure variable which was living on a farm. There is plausibility to this result, as direct contact with animals has been previously identified as an important risk factor for cryptosporidiosis (62–64), but this is often outbreaks and not sporadic disease.

The Tollestrup (a) exposure measured direct handling of livestock and the study reported reduced odds. Although curious, this could be due to repeat or continuous exposure to the animals producing an elevated serological response in some participants. The use of a long-term marker, such as 27-kDa, as an outcome for demonstrating *Cryptosporidium* infection, has an impact on the interpretation of results.

#### Pets/Domestic animals

Two studies found significant positive associations with owning or handling pets. (Fournet (2), OR=2.56; 95% CI=2.3-2.84 and Tollestrup (b), OR=2.83; 95% CI=1.24-6.29). The Tollestrup (c) study did also investigate this exposure and it remained in the model; we have not reported it here with the other studies as it was related to the 15/17-kDa marker, and our results reflect positivity using 27-kDa. (**Error! Reference source not found**.)

Specifically, the Fournet (2) study considered dog ownership and as most of the cases were *C. hominis*, a more anthroponotic species, this is a peculiar result. However, we know that in this study exposures for cases were compared between years, and it may be a chance effect that dog ownership in the population changed, or indeed that it is a proxy exposure for an uncaptured variable.

These results are unusual: little epidemiological evidence exists to suggest pets have any role to play in the transmission of disease (52,65,66) especially for the two main species. The quality and robustness of these two studies do not allow complete confidence in the results: the Fournet (2) study reported on cases between time periods, and the Tollestrup (b) study used serological responses as a marker for infection and so must be interpreted differently to current disease. Yet, there is some evidence that oocyst shedding in cats and dogs can occur (26,67) and species-specific infections have, albeit rarely, been detected in humans (68,69).

This theory requires further investigation to be fully understood and described, and the addition of species identification in any study of this exposure may help better understand the true risks and pattern of infections in both domestic pets and their owners.

### Transmission pathway - Food exposures

Results for food exposures were reported in three of the included studies (four variables) and this was the least investigated exposure route when considering all of the studies’ upfront designs. All found significant results. **Error! Reference source not found**. shows results with the specific metric included: due to the variability of measures and the small number of results reported in the pathway, we did not separate exposures. Two variables (eating from a farm stand and eating tomatoes) were associated with reduced odds of illness (Valderrama; OR=0.2, 95% CI=0.1-0.9; and Nic Lochlainn; OR=0.6, 95% CI=0.5-0.8).

Two variables (eating BBQ food and eating outside the home) were associated with an increased risk of illness (Nic Lochlainn; OR=1.8, 95% CI=1.4-2.3; and Fournet (UK); OR=11.32, 95% CI=9.36-13.7). The Pintar study did study several food variables appropriate to eating outside the home, but none remained in the final models, likely due to the limitations of using surveillance data (50).

Previous work in Europe has increasingly demonstrated risks associated with food (15,18,70–72). Our results indicate a range of risks, but exposures measured were specific to each study and its setting, making comparisons and assessment overall difficult. However, ‘eating tomatoes’ is consistent with the literature where raw, salad food item appear to be associated with reduced disease risk, such as carrots (73) and tomatoes (66)

Eating outside the home is often a risk factor for any infectious intestinal disease but in the case of *Cryptosporidium* might be confounded by multiple factors. The profile of food-borne disease is shifting, affected by globalisation and changes in food practices (74). Additionally, advances in diagnostic methods and surveillance systems have extended the range of protozoa that may be linked to food (70) and this exposure should be further explored (75,76).

### Transmission pathway - Person-to-person

Person-to-person, as a transmission pathway, was well investigated overall (five studies, four included) and represents the most consistent finding so far.

#### Contact with a case of diarrhoea

All three studies reporting on person-to-person contact with a symptomatic individual demonstrated correlations between exposure and disease (Pintar: OR=2.86; 95% CI=1.28-6.38 and OR=2.1, 95% CI=0.64-6.87; Nic Lochlainn: OR=2.2; 95% CI=1.7-3.0; and de Gooyer: OR=12.6; 95% CI=2.13-75.1).

Variables included both home and outside-the-home contact, perhaps demonstrating the importance of differences between case contact in a household where caring and close contact is more likely and contact in a non-shared space. The de Gooyer and Nic Lochlainn studies, and one Pintar variable, all related to household transmission. The Pintar non-household contact variable reported lower odds and wider confidence intervals crossing 1.0, suggesting that transmission within the home is more important as a risk factor for sporadic disease. Additionally, this study excluded cases that initially had reported other illness in the home in order to accurately identify community index cases. This would suggest that any known cases came after the index illness, indicating that onward spread is more prevalent in *Cryptosporidium* than the other enteric illnesses.

Contact with a case is a well-known, and plausible, transmission pathway to disease and exposures underlying this pathway are varied including childcare, and sexual transmission (Hannah and Riordan, 1988; Hellard *et al*., 2003; Hunter *et al*., 2004; Artieda *et al*., 2012; Johansen *et al*., 2014). This makes biological sense given the faecal-oral route of transmission and the high prevalence in younger children who may require help with toileting. The fairly high odds ratios demonstrate the importance of this pathway to disease, particularly in the home environment. The included studies here have investigated symptomatic contact, but asymptomatic carriage has also been identified as a possible factor in transmission of sporadic disease in the home environment (22,23,66).

#### General contact

Variables which investigated social contact without the prerequisite of contact with a symptomatic individual, were classed under general social contact and ranged from quite specific, e.g. child-care, to broad, e.g. any social contact. All of the results were statistically significant.

Two studies showed a decreased risk of disease associated with general person-to-person contact measured as attendance at ‘any social gathering/event’ (Pintar: OR=0.23; 95% CI=0.05-0.99) and (Valderrama: OR=0.4, 95% CI=0.1-0.9).

Both of these variables were investigating general social contact and might cover a range of activities and undoubtedly have a high exposure in all groups, thus the result is likely to be spurious.

The Valderrama, 2009 study explored ‘contact with a child in diapers’ among community cases of disease and found an almost four-fold risk of disease (OR=3.8, 95% CI=1.5-9.6). This could be driven by the higher prevalence observed in younger children, as well as the possible contribution of asymptomatic spread and is probably best considered separately to ‘general social contact’. It has also been demonstrated that young children are drivers of secondary spread of disease, whether they are symptomatic or not (66).

Considering the person-to-person exposures together, it seems reasonable to suggest that the contribution of cases to onward spread warrants further investigation. This seems to apply particularly to the home environment, where we might identify easy and meaningful public health interventions to mitigate spread (79). Additionally, when symptoms are used to define a case, we might be losing vital information on asymptomatic disease, and how much that contributes to spread of infection.

### Transmission pathway - Water

Water exposures were investigated in nine of the ten included studies. This pathway was further disaggregated into drinking and recreational water exposures, with nine and five studies investigating these, respectively.

#### Drinking water

This was categorised as treated (n=2); untreated (n=5); and bottled (n=3). (Figures 8-10)

##### Treated

One study (Pintar, 2009) found a significant result between consumption of municipal treated drinking water and an increased risk of disease (OR=2.43; 95% CI=1.0-5.7).

There had been a sharp increase in cases of *Cryptosporidium* in this area just prior to the study period. It is possible that the association between municipal war supplies was due to a specific contamination and perhaps represents an outbreak, or was a proxy for another association, such as rural versus urban residence. The other study (Fournet, (1) 2013 shows an insignificant decreased risk of disease (OR=0.56; 95% CI=0.29-1.05), with a smaller sample size. The metric was, “drank tap water on daily basis” which is likely a prevalent and difficult to accurately measure metric. This study was based on an observed increase in cases and comparisons were made between years – it is feasible that drinking treated water may appear to have a protective effect in comparison to drinking from an untreated or contaminated water source, particularly if the increase was actually an undetected outbreak. Additionally, the odds ratios for the Fournet, (1) study were calculated using presented data and are not those reported in the manuscript. As such we are unable to comment on any confounders or control for effects.

An interesting finding here is that although water exposures were commonly investigated, examinations were not often directed at drinking water, and more often studies considered variables related specifically to drinking from untreated sources. Although drinking water is often the cause of outbreaks and is considered to be the main point for public health intervention only two of our studies reported final results allocated to this pathway, and both studies were following an undetermined increase in cases, which may not truly represent sporadic disease. It may be that following recent water treatment and regulatory requirements this is now considered a less burdensome transmission pathway for disease (80).

##### Untreated

Two studies reported significant positive effects between the consumption of untreated drinking water and odds of infection with *Cryptosporidium*, ranging from almost two-fold (Tollestrup (a): OR=1.98; 95% CI =1.11-3.55) to an increased risk of eight times that of controls (Valderrama: OR=8.0; 95% CI=1.3-48.1).

The 2009 Valderrama study had wide confidence intervals, the lower end of which was close to 1.0, despite quite a high odds ratio (8-fold). The data analysed were based on surveillance data extractions and so there are possible limitations on questions asked and answered. The authors grouped all and any untreated water consumption. Additionally, although not reported as an outbreak, this study was in response to an increase in cases across the state of Colorado, USA, and it may be possible that the elevated risks are a result of an outbreak driven by a specific untreated water source during the study period, although controls were matched by geography which should mitigate this to some extent.

The Tollestrup (a) study included 200 of the 600 cohort participants, taking place in a semi-rural area, where there was a significantly higher percentage of participants using onsite wastewater systems or private wells than participants using municipal systems (p=0.048). It is also important to consider that the outcome measured in this study was serological response to 27kDa antigen rather than diarrhoeal illness and so our confidence in comparability may be reduced. Ongoing non-transient exposure to oocysts, perhaps in an untreated system, could result in a positive, and increasingly high, serological response in residents in this area. This may not, however, necessarily reflect current or prior disease.

The remaining non-significant results were all fairly close to 1.0 with narrow confidence intervals. Results from the Becker (2015) study were not significant and close to no effect (aOR=1.19; 95% CI=0.97 - 1.45), despite being a large study of high quality. As with the Tollestrup studies, the outcome metric was serological response to a *Cryptosporidium* marker, which might indicate differences in exposure windows and actual infection cannot be accurately pinpointed.

It is interesting to see that this is a well investigated pathway, and it is biologically plausible that exposure to untreated drinking water could represent a source of *Cryptosporidium* oocysts. Nevertheless, even with the studies’ limitations in mind, these results do not seem to indicate that this pathway is a major contributor to sporadic disease.

##### Bottled

All of the studies including results for this pathway reported positive associations with disease. Two of the three studies found a significant positive effect between drinking bottled water and increased odds of *Cryptosporidium* infection, ranging from almost three-fold (Fournet NL (1), 2013: OR=2.72, 95% CI=1.1-6.76) to more than five-fold (De Gooyer, 2017: OR=6.31; 95% CI=1.39-28.7).

The odds ratio for the Fournet (1) study in the Netherlands was calculated using their reported data: They reported a 21% vs 11% exposure in cases and controls respectively. Most cases in the study were infected with *C. hominis* (GP60 subtype IbA10G2) which usually suggests either anthroponotic spread or perhaps sewage contamination.

The de Gooyer study investigated sporadic cases in Australia, where the consumption of bottled water in the general proportion is fairly high (48) and, in this study, 86% of cases vs 54% of controls exposed was reported. Although exposure prevalence in the control group was lower, it still represents a reasonable amount of non-diseased participants that cannot be explained by that exposure.

The Fournet (2) UK study reported narrow confidence intervals yet did not demonstrated a difference in effect. The controls in this study were cases from different years: it is reasonable that drinking bottled water may have no effect on disease incidence in comparison to a period of time where cases were driven by an undetected outbreak with a single other source.

If exposure to bottled water was a risk for sporadic disease (and not outbreaks following a specific contamination) we might expect to see exposure to tap water generally associated with a decreased risk of illness in those exposed, but two case-control studies in areas close to the de Gooyer investigation in Australia demonstrated no such effects of water associated with *Cryptosporidium* (65). Also, paradoxically, water consumption consistently free from *Cryptosporidium* oocysts may be associated with reduced immunity over time potentially increasing susceptibility to infection (81) but this is still poorly understood (32,82). However, *C. hominis* has been detected in finished mineral water samples following an outbreak in the UK (83) and Australia (84) suggesting some plausibility in these results. In these studies, definitions of ‘bottled water’ varied and information wasn’t specifically collected on amounts consumed to enable further examination of this association. Additionally, sociodemographic factors associated with bottled water use have previously been described (85) and so there is a possibility that these results might be open to uncontrolled confounding.

On balance, there is some evidence for bottled water as a risk for sporadic illness, but these studies offer insufficient quality and detail to make a resolute conclusion.

#### Recreational water

This was categorised as treated (n=3); and untreated (n=3). (Figures 11 & 12)

##### Treated

Three studies reported on exposures that were categorised as treated recreational water, and all found significant results. Two studies reported an increased risk of disease associated with exposure to recreational water (Valderrama, 2009: OR=4.6, 95% CI=1.4-14.6; de Gooyer, 2017: OR=26.4, 95% CI=1.47–472 for general spa use).

The de Gooyer study investigated two variables related to recreational water exposure: using a spa and having any recreational water exposure. These were wide ranging definitions and captured any and all recreational water exposures (except those that might be specifically described as untreated). As such, we cannot be certain that specific exposures in this group are necessarily treated waters. This could misclassify the exposure and overestimate the effect. Using a spa was significantly associated with illness (p=0.03), although only 13% (4/30) of cases reported this exposure. We also know that this work was examining an increase in cases that may have been linked to a particular recreational waterpark, although was not considered an outbreak. If these cases were linked to a particular water source, this could skew the results towards a positive effect.

It is worth noting that in Australia, recreational water exposures are a common cause of outbreaks and the prevalence of this recreational activity is high (86,87). These activities represent a biologically plausible route to infection with *Cryptosporidium*, considering its faecal-oral transmission route and the likelihood of swallowing water, the chlorine resistant nature of oocysts, and the poorer hygiene habits of younger children (12).

One of the studies reported decreased odds of disease (Fournet UK, 2013: OR=0.37, 95% CI=0.33-0.42). This study originally reported proportions and we used these data to calculate odds ratios – thus these are not adjusted or controlled for any other factors, which may skew the effect measure. Also, the study is considering cases from one time period versus another, rather than comparing exposures between diseased and non-diseased participants. It is reasonable that the reported effect could be an artefact if recreational water exposure was the cause of the increase in the prior years. Whilst we may be able to confidently say that exposure to treated recreational water was not causing the increase in the study time period, we can be less confident that the results are generalisable and that they represent the true relationship between exposure and sporadic disease.

It is likely that treated recreational water represents some risk for cryptosporidiosis, and this would be supported in other literature. However, these results suggest that it may well be more associated with outbreaks, or specific incidences of increases in disease.

##### Untreated

One of three studies reporting on untreated recreational water exposures found a significant result, demonstrating a three-fold increased odds (Pintar, 2009; OR=2.91, 95% CI=1.14-7.38). Figure 12

After adjusting for age and season in the multivariable model, swimming in an untreated water venue (river or lake) (p=0.01) was associated with increased odds of illness. The proportion of cases exposed was, however, small at 25% (9/36) versus 10% (79/801) for the controls (79/801). This study used other cases of enteric illness as controls, including *Giardia*, which has similar exposure routes. This could make it more difficult to detect small differences in risk between case and controls by over-representing exposures and biasing towards the null, so in actual fact this effect measure could be underestimated (88). The data were however, taken from surveillance records, and cases were only asked about exposures in the seven days preceding illness which might not accurately capture the exposure window for *Cryptosporidium*.

The remaining two studies were statistically insignificant, and both reported no effect of this exposure on risk of disease in their study population (Fournet (1) NL (OR=1.15, 95% CI=0.63-2.1) and de Gooyer (OR=1.0, 95% CI=0.31-3.25).

Overall, the contribution of this exposure to disease in this review is not considerable. This is interesting in light of the more positive effects we report for treated recreational water. This may be due to untreated activities being associated with other unmeasurable factors, such as adult, rather than child, populations (89).

### Publication Bias

Due to the small number of studies against each exposure category (n<5), we did not consider it appropriate to proceed with statistical analysis into heterogeneity and publication bias.

## Conclusions

The case-characteristics collected from our included studies reflected the bimodal age-related pattern that *Cryptosporidium* follows in the US and the UK; a peak in young children, and a peak in adulthood (21,90,91).

The animal transmission pathway was commonly investigated, although was not considered a main hypothesis for sporadic disease in any of the papers. We report a couple of unusual associations with disease and pet contact, specifically cats and dogs, although these should be considered in light of some of the methodological weaknesses of the studies concerned. Evidence for this route is conflicted and more research is needed to support or refute this pathway as a contributor to sporadic infection.

Food exposures were not so frequently investigated, and metrics used were specific to the niche study population or hypothesis. Given that food items are increasingly identified in outbreak investigations (Casemore, 2001; Ethelberg *et al*., 2005; McKerr *et al*., 2015). this exposure, and protozoa that may be linked to food, should be further explored (70,75,76).

Person-to-person transmission was well investigated overall and represents the most consistent finding so far. Considering the person-to-person exposures, it seems reasonable to suggest that the contribution of cases to onward spread warrants further investigation. This seems to apply particularly to the home environment which is increasingly understood to be a significant setting for spread of *Cryptosporidium* infection (22,23,79,92) and would support public health messaging on preventing spread of disease at home (93).

Our included papers investigated water exposures most frequently despite evidence that in industrialised countries in recent years, drinking treated mains water is unlikely to cause a significant amount of sporadic cryptosporidiosis (94,95). However, despite improved legislation for drinking water quality (80) deficiencies can persist and it is important to keep monitoring this pathway (96,97). We present some risks associated with bottled water, although this is likely to be a high prevalence exposure and may be confounded by socio-economic variables.

Recreational water is more frequently associated with outbreaks and was not a major source of sporadic illness in this review, perhaps reflecting the episodic nature of pool water contamination events. Nonetheless, we know that standard treatment practices for recreational water, such as chlorination, are unsuccessful in eliminating *Cryptosporidium* oocysts (98). This does seem a well understood pathway (2,99).

When considering exposures for sporadic disease, it may be more pertinent for future studies to focus on food exposures, and research in this area is on the increase (100).

Secondly, our results demonstrate that further detail is required on the case-to-person transmission pathway: although well investigated, exposures were variable, and no study hypothesised this as a risk for infection, with most results incidental to the study. We should seek to quantify and ascertain spread of infection in the home environment and build a profile of asymptomatic infections, through better observational studies and more routine sub-typing of isolates (101,102).

## Limitations

As with any work which seeks to combine various different studies, significant limitations exist which should be considered in any synthesis.

The main limitations to this work were the low number of papers included, which meant meta-analysis was not possible. Although we didn’t quantify heterogeneity, anecdotally, there were differences between the studies included in terms of study design, populations studied, data collected and variables measured, and outcomes, which may have made meta-analyses difficult regardless.

In the studies, often several variables were measured which represented the same exposure, and participants may have been counted in either as they were not necessarily mutually exclusive. This is common to epidemiological studies, which are often undertaken in response to outbreaks, and defined by particular settings and putative exposures (29). The breadth of this makes the extrapolation of our results less robust, and context must be considered in assessing results. This might dilute any differences between exposures driving sporadic disease and outbreaks. A suggestion for further work would be to extend the time period to look specifically at the magnitude of changes in exposures and to review outbreaks in that context. Unfortunately, this was beyond the scope of this work.

Additionally, there could be bias associated with the personal subjectivity of categorising exposures into pathways, especially without the granular detail of raw data or knowing the specifics of questions asked of participants. However, our robust methodological approach and commitment to duplicating all tasks in this review hopefully mitigates this as far as possible.

Two included papers (45,51) measured their outcome using serological response to the 15/17kDa an 27kDA antigen groups, which can identify previous, as well as recent infection and cannot distinguish between species or genotypes (103). Because we had two papers with this outcome, which judged it differently, we used response to the 27kDa to indicate a strong serological response and therefore count as infection. However, this has a longer positivity than the 15/17kDa, and could represent infection as long as nine months prior (53). This might have meant we were correlating current or recent exposures to old infection and making inferences when in fact there was no link. However, as we were looking at sporadic infection as opposed to outbreak disease, it may be appropriate to consider more static exposures, in any case. Additionally, diarrhoeal disease from *Cryptosporidium* infection may well recur and has comparative longevity, that might well underpin some health seeking behaviour, so the other studies are also at some small risk of correlating exposures with disease in a different time frame. Furthermore, even those studies that do have accurate and recent onsets are often a) vulnerable to recall biases and b) have different windows of exposure for cases and controls (27).

Often crucial detail was absent from the manuscript, and so allocation of measured variables to our exposures was occasionally arbitrary. However, we moderated this as far as possible by following a systematic approach and utilising our third reviewer for any discord. We would recommend that future research includes large-scale observational studies with enough resource to achieve high study quality whilst maintaining resolution in exposures measured.

In assessing study quality, we recognised that the NOS has significant limitations for use in this type of systematic review (104). A number of studies we reviewed used surveillance data rather than more traditional study designs which meant that the NOS was not always directly applicable. Where this was the case, we have had to apply the tool as appropriately as possible and in discussion with reviewers. At present, there is no risk of bias tool which encompasses both case-control studies and surveillance data.

Despite the limitations considered, this is the first systematic review considering routes of transmission in industrialised countries for sporadic *Cryptosporidium* and highlights that food routes are under investigated, and that person-person transmission, although recognised, is not thoroughly investigated.

## Supporting information

PRISMA Flowchart

PRISMA Checklist

## Data Availability

All data produced in the present study are available upon reasonable request to the authors

## Abbreviations

AIDS: Acquired Immunodeficiency Syndrome
GRADE: Grading of Recommendations, Assessments, Development and Evaluation
HIV: Human Immunodeficiency Virus
NOS: Newcastle-Ottawa Scale
NIHR: National Institute for Health Research Health Protection Research Unit
OECD: Organisation for Economic Co-operation and Development
OR: Odds Ratio
PHE: Public Health England
PHW: Public Health Wales
PRISMA–P: Preferred Reporting Items for Systematic Reviews and Meta-Analyses for Protocols
ROBINS-I: Risk of Bias In Non-randomized Studies - of Interventions
RR: Relative Risk/Risk Ratio
UK: United Kingdom

## Declarations

Nothing to declare.

## Ethical approval

As this is a literature review, ethical approval is not indicated or required. Research ethics approval is not required for research involving information freely available in the public domain.

## Competing interests

The authors declare that they have no competing interests.

## Funding

This research is jointly funded by the National Institute for Health Research Health Protection Research Unit (NIHR HPRU) in Emerging and Zoonotic Infections (HPRU EZI) and the National Institute for Health Research Health Protection Research Unit (NIHR HPRU) in Gastrointestinal Infections (HPRU GI) at the University of Liverpool in partnership with Public Health England (PHE), University of East Anglia, University of Oxford and the Institute of Food Research. The views expressed are those of the authors and not necessarily those of the NHS, the NIHR, the Department of Health, PHE, or Public Health Wales.

## Authors’ contributions

**CMCK, RChr, RCha, RV, and SOB** conceived the initial idea for the study.

**CMCK**: Paper sourcing, Extraction & Analysis, Writing – original draft, Writing – review & editing, Data curation

**WS:** Data extraction tools, Extraction & Analysis, Writing – original draft, Writing – review & editing

**AW:** Title screening and first round inclusion of manuscripts, protocol review

**KGP:** Third reviewer, manuscript writing, critical revision

All authors read and approved the final manuscript.

## Acknowledgements

Thanks to Ken Linkman at the University of Liverpool Harold Cohen library for his specialist aid with medical databases and search terms, Dan Pope at University of Liverpool Public Health Department for technical guidance for data synthesis, and to Peter Burrell at Public Health England for assistance with data extraction tools.

Erica Kintz at University of East Anglia and Ruaraidh Hill at Liverpool Reviews and Implementation Group (LRiG), University of Liverpool for advice on design and data collection.

## Tables and Figures

Table 1: Newcastle-Ottawa Scale scores with individual domains and general strengths and weakness of the study design

Table 2: General characteristics of included studies

Table 3: Characteristics and main exposures or factors measured, and main results.

Figure 1: PRISMA diagram showing manuscript capture and inclusion

Figure 2: Transmission pathways and underlying exposures used to categorise variables measured in the included studies

Figure 3: Exposure measured and results: Animal contact – Farm animals

Figure 4: Exposure measured and results: Animal contact – Pets

Figure 5: Exposure measured and results: Food

Figure 6: Exposure measured and results: Person-to-Person - Contact with a case of diarrhoea

Figure 7: Exposure measured and results: Person-to-person – General contact

Figure 8: Exposure measured and results: Drinking Water - Treated

Figure 9: Exposure measured and results: Drinking Water – Untreated

Figure 10: Exposure measured and results: Drinking Water - Bottled

Figure 11: Exposure measured and results: Recreational water – Treated

Figure 12: Exposure measured and results: Recreational water – Untreated

## Additional/Supplementary files

Additional file 1: OECD membership

Additional file 2: Search terms

Additional file 3: Minimum data set for collection

## Notes

### Competing Interest Statement

The authors have declared no competing interest.

### Clinical Protocols

https://www.crd.york.ac.uk/prospero/display_record.php?RecordID=56589

