## Supplementary material for "Exposures associated with sporadic *Cryptosporidium* infection in industrialised countries: a systematic review": Data Figures

Figure 2: Transmission pathways and underlying exposures used to categorise variables measured in the included studies

Animals

Pets

Farm animals

Food

Person-to-person

Close contact

General social contact

Water

Drinking water

Treated

Untreated

Bottled

Recreational water

Treated

Untreated

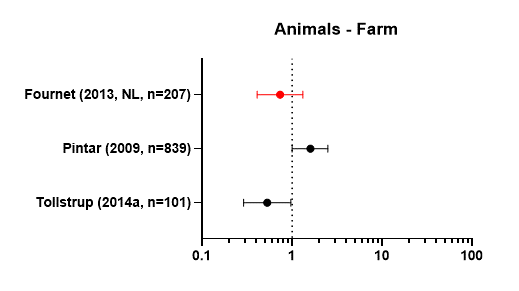

*Figure 3: Exposure measured and results: Animal contact – Farm animals*

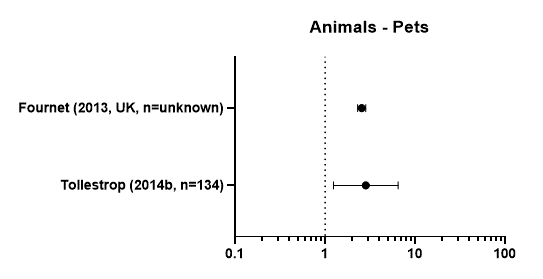

*Figure 4: Exposure measured and results: Animal contact – Pets*

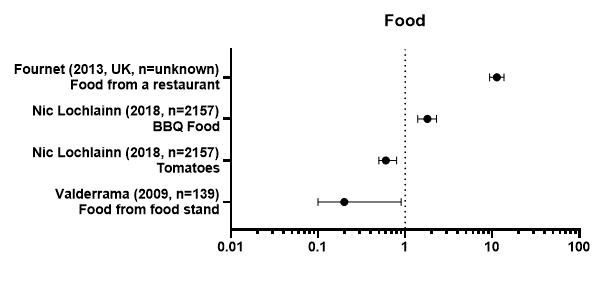

*Figure 5: Exposure measured and results: Food*

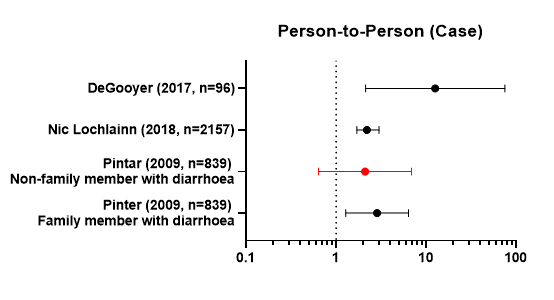

*Figure 6: Exposure measured and results: Person-to-Person - Contact with a case of diarrhoea*

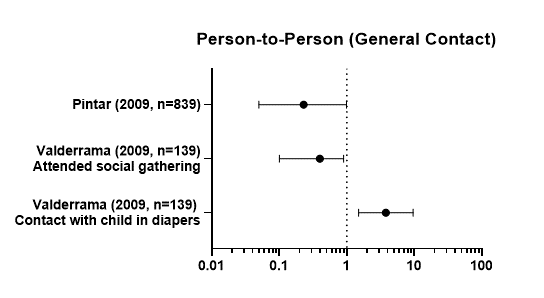

*Figure 7: Exposure measured and results: Person-to-person – General contact*

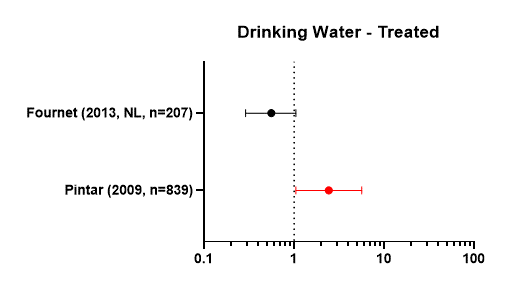

*Figure 8: Exposure measured and results: Drinking Water - Treated*

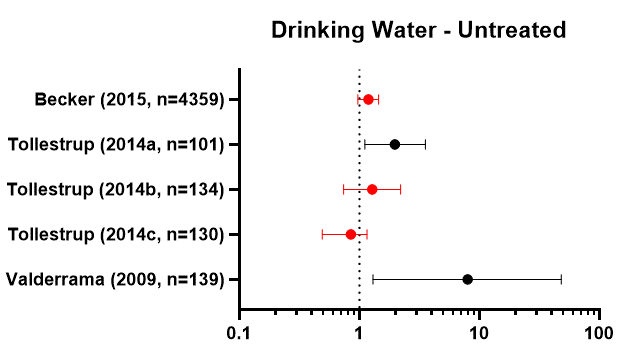

*Figure 9: Exposure measured and results: Drinking Water – Untreated*

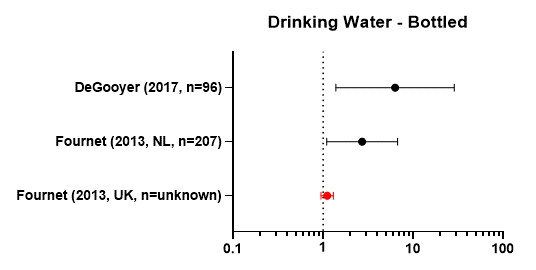

*Figure 10: Exposure measured and results: Drinking Water - Bottled*

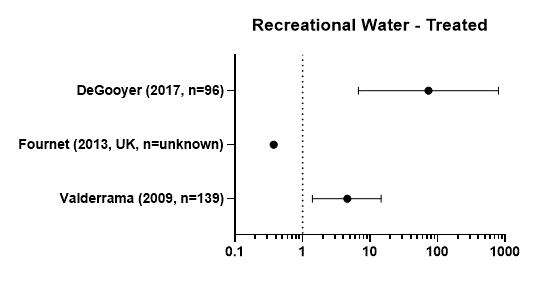

*Figure 11: Exposure measured and results: Recreational water – Treated*

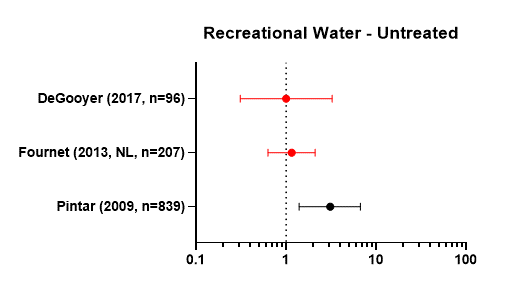

*Figure 12: Exposure measured and results: Recreational water – Untreated*
