## Supplementary material for "Exposures associated with sporadic *Cryptosporidium* infection in industrialised countries: a systematic review": Data Tables

Table 1: Newcastle-Ottawa Scale scores with individual domains and general strengths and weakness of the study design

|  |  | |  |  | **Selection** | | | | **Comparability** of Cases & Controls | **Outcome** | | |
| --- | --- | --- | --- | --- | --- | --- | --- | --- | --- | --- | --- | --- |
| **Study design** | **Author/Year** | **Overall NOS Score (quality)** | **Strengths** | **Weaknesses** | Case Definition | Case Representativeness | Selection of Controls | Definition of Controls |  | Exposure Ascertainment | Case/Control Ascertainment | Non-Response Rate |
| Cross-sectional | Ravel (2012) | **6**  **(moderate)** | - Surveillance data provided  a rich data set which can be accessed with minimal resource burden  - Multiple exposures could be investigated  - Authors were able to clearly exclude known outbreak-related cases | - Surveillance data can bias participation as only included captured cases  - Relevant to a specific area, which may decrease generalisability of results  - Compared cases to other enteric pathogens, so the exposure results are relative to other IID (with similar pathways) | * |  | * | * | * | ***** | ***** |  |
| Case-control | NicLochlainn (2018) | **5**  **(moderate)** | - Timely administration of questionnaire so recall of past exposure(s) may be more accurate  - Laboratory diagnosed so less chance of misclassification of cases  - Large-scale and representative | - Surveillance data can bias participation as only includes captured cases  - Exposures reported over 3 years, separately but lack of raw data means we could only use the final model  - Possibility of selection bias among those who completed and returned the exposure questionnaire | * |  | * | * | * |  | * |  |
| Case-control | De Gooyer (2017) | **5 (moderate)** | - Frequency matching allows control for any confounding role of age  - Had a clear hypothesis for exposure and outcome  - Collect exposure data 14 days prior to illness, comparable with *Cryptosporidium* exposure window | - Small numbers affects power and representativeness of results  - Followed an increase in cases that was not classed an outbreak, but may bias results towards a point source  - Controls were contacted in a different time period to cases which may introduce recall biases | * |  | * | * | * |  | * |  |
| Case-control | Fournet (1) (2013) | **7**  **(high)** | - Controls randomly selected and representative  - Good control to case ratio  - Matching allows control for any confounding role of age and sex | - Followed an increase in cases that was not classed an outbreak, but may bias results towards a point source | * |  | * | * | * | * | * | * |
| Case-control | Valderrama (2009) | **5**  **(moderate)** | - Two controls per case  - Matching allows control for any confounding role of age and geography  - Controls randomly selected and representative | - Low response rate  - Small numbers  - Followed an increase in cases that was not classed an outbreak, but may bias results towards a point source  - Controls were contacted in a different time period to cases which may introduce recall biases | * |  | * | * | * |  | * |  |
| Case-case | Fournet (2) (2013) | **4**  **(low)** | - Surveillance data provides  a rich data set which can be accessed with minimal resource burden  - Multiple exposures could be investigated | - Followed an increase in cases that was not classed an outbreak, but may bias results towards a point source  - Compared current cases to case in previous years, so identified exposures may reflect an undetected outbreak or single source  - The case-case study was in a small area and the authors did not report numbers so results may not be generalisable | * |  |  |  |  | * | * | * |
| Case-case | Pintar  (2009) | **5**  **(moderate)** | - Looked at a reasonably long time period  - Authors were able to exclude travel and outbreak cases  - Laboratory diagnosed so less chance of misclassification of cases | - Compared cases to other enteric pathogens, so the exposure results are relative to other IID (with similar pathways)  - Used smaller surrogate models due to low power | * |  | * | * | * | * | * |  |
| Serological | Becker  (2015) | **6**  **(high)** | - Used a well-established, representative and systematically collected dataset (NHANES)  - Case was defined as a positive IgG response to both markers, strengthening the case definition  - Large numbers | - Restricted to ages 6-49 which might exclude key age groups  - This type of large-scale data collection might bias towards a certain characteristic(s) or responder  - General methodological issues with serology studies for *Cryptosporidium*, including;   - Increasing response with age - Hard to measure impact of non-transient exposures - Not clear when infection or disease occurred so difficult to establish accurate exposure window - Cannot distinguish species   - Authors report mostly on characteristics of cases and socioeconomic factors rather than exposures | * | * | * | * | * |  | * |  |
| Serological | Tollestrup (2014) | **4**  **(low)** | - At least one study ran for more than a year  - Good study size  - Able to collect environmental data and blood samples  - Multiple exposures could be investigated | - Restricted to adults only which might exclude key  - General methodological issues with serology studies for *Cryptosporidium*, including:   - Increasing response with age - Hard to measure impact of non-transient exposures   - Cannot establish any temporal association between exposure and outcome - 27-kDa marker may reflect chronic, long-term exposure(s)  - Environmental samples collected after exposure  - Convenience sampling may introduce selection bias |  |  | * | * | * |  | * |  |

Table 2: General characteristics of included studies

| Study reference | Study year(s) | Country | Design | Hypothesis/  main exposures investigated | Recruitment/Identification of cases | Outcome measured | Cases / controls | Quality |
| --- | --- | --- | --- | --- | --- | --- | --- | --- |
| Ravel *et al*  (46) | 2005-2009 | Canada | Cross-sectional | Clinical, demographic and exposure variables associated with sporadic, domestically acquired parasitic disease. Cases were assigned transmission routes based on: animal-person, environment, water, food, person-person routes | National surveillance dataset | Laboratory confirmed *Cryptosporidium* excluding known foreign travel acquired and outbreak cases | 176 | Moderate |
| NicLochlainn *et al*  (47) | 2013-16 | Netherlands | Case-control | General risk factors for sporadic cryptosporidiosis: demographics, symptoms, medical history, foreign travel, contact with animals, contact with ill persons, recreational activities, and food and drink consumption | National surveillance dataset | Laboratory confirmed *Cryptosporidium* with gastrointestinal illness (controls were selected from resident population and free from diarrhoeal illness) excluding foreign travel | 609 / 1,548 | Moderate |
| de Gooyer *et al*  (48) | February - March 2015 | Australia | Case-control | To identify risk factors associated with region-wide increase in cryptosporidiosis with specific focus on recreational water activities as a hypothesis | National surveillance dataset | Laboratory confirmed *Cryptosporidium* cases under 50 years old (controls were asymptomatic resident population) | 30 / 66 | Moderate |
| Fournet *et al* (1) NL  (44) | 2012 (late summer) | The Netherlands | Case-control | To identify risk factors for an increase in sporadic cases compared to previous years, with a focus on accepted transmission routes: farm animals, recreational water, drinking water, including bottled, and travel history | National surveillance dataset | Laboratory confirmed *Cryptosporidium* case with diarrhoeal symptoms (controls were randomly selected from resident population) | 82 / 125 | Moderate |
| Valderrama *et al*  (49) | August -September 2007 | USA | Case-control | To identify risk factors for an increase in sporadic summer cases, specifically for community disease. Exposures included food and water consumption, recreational water, childcare, animal contact, person-to-person contact, and travel history | National surveillance dataset | Laboratory confirmed *Cryptosporidium* (controls were asymptomatic l resident population) | 47 / 92 | Moderate |
| Fournet *et al* (2) UK  (44) | weeks 32 to 42 of 2012 | UK (North-East region) | Case-case | To identify risk factors for sporadic cases compared to previous years, with a focus on accepted transmission routes: farm animals, recreational water, drinking water, including bottled, and travel history | National surveillance dataset | Laboratory confirmed *Cryptosporidium* (controls were cases from previous 3 years) | 3,230 / 3,281 for whole of UK. Unreported numbers for the case-case study in the North East region. | Moderate |
| Pintar *et al*  (50) | 2005 - 2007 | Canada | Case-case | Determining exposures for sporadic, endemic disease - addressed three routes of exposure based on *a priori* hypotheses: recreational water, environ-mental, and person-to-person transmission | National surveillance dataset | Laboratory confirmed *Cryptosporidium* excluding known foreign travel and outbreak cases (controls were cases of other gastrointestinal pathogens) | 36 / 803 | Moderate |
| Becker *et al*  (51) | 1999-2000 | USA | Serological study (with cross-sectional approach) | Investigated correlates of social inequality (food adequacy, annual income, and the poverty income ratio) against the odds of positive serological response | Large-scale national community health/surveillance database (NHANES) | Positive IgG response to both the 15/17kDa and 27kDA *Cryptosporidium* antigens in sera, subjects aged 6-49 years old | 925 / 3,434 | Moderate |
| Tollestrup *et al* *(a,b,c)*  (3 studies at separate sites)  (45) | Study sites:  a: 2004 - 2005  b: August 2006-February 2007  c: May - October 2006 | USA | Serological study (with cohort approach) | Investigated if wastewater from onsite systems and private water supplies correlated to serological response: Hypothesis: that participants living in households with onsite wastewater systems and private wells would be more likely to have elevated serological responses compared to those living in households using municipal systems because of increased exposure to *Cryptosporidium o*ocysts. | Community recruitment and provision of a blood sample | Positive IgG responses to the 15/17-kDa and 27kDa *Cryptosporidium* antigens in sera in >18-year olds | 600 | Low |

Table 3: Characteristics and main exposures or factors measured, and main results.

| **Study type** | **Paper/Study** | **Country** | **Transmission pathway/risk factor** | **Exposure measured** | **^[1]^Measure of association (reported)** | **95% CI** | **p** | **Level** |
| --- | --- | --- | --- | --- | --- | --- | --- | --- |
|  |  |  |  |  |  |  |  | **of analysis** |
| **Cross-sectional** | **Ravel, 2013** | **Canada** | Animals | Contact with household pets | Proportion exposed = 49 | 34-64 | - |  |
|  |  |  | Animals | Visited a farm, a petting zoo or fair | Proportion exposed = 18 | 9-32 | - |  |
|  |  |  | Food | Ate food prepared outside home | Proportion exposed = 43 | 28-58 | - |  |
|  |  |  | Food | Ate meat from any place other than the grocery store | Proportion exposed = 26 | 14-40 | - |  |
|  |  |  | Food | Drank or ate any unpasteurised milk, juice, or dairy products | Proportion exposed = 8 | 2-19 | - |  |
|  |  |  | Food | Shopped at a supermarket | Proportion exposed = 93 | 82-99 | - |  |
|  |  |  | Food | Shopped at butcher shop | Proportion exposed = 20 | 9-34 | - |  |
|  |  |  | Food | Shopped at farm | Proportion exposed = 7 | 1-18 | - |  |
|  |  |  | Food | Shopped at farmer’s market | Proportion exposed = 4 | 1-15 | - |  |
|  |  |  | Outdoor activities/environmental exposure | Canoed, kayaked, hiked, or camped | Proportion exposed = 16 | 7-30 | - |  |
|  |  |  | Outdoor activities/environmental exposure | Gardening | Proportion exposed = 13 | 5-26 | - |  |
|  |  |  | Outdoor activities/environmental exposure | Lived on a farm or country property | Proportion exposed = 30 | 18-45 | - |  |
|  |  |  | Person to person | Attended social gatherings | Proportion exposed = 18 | 9-32 | - |  |
|  |  |  | Person to person | Hosted or attended a barbeque | Proportion exposed = 33 | 20-49 | - |  |
|  |  |  | Person to person | Knew anyone outside the household with a diarrhoeal illness | Proportion exposed = 13 | 5-25 | - |  |
|  |  |  | Travel | Domestic travel | Proportion exposed = 100 | 69-100 | - |  |
|  |  |  | Water | Bottled water | Proportion exposed = 53 | 39-67 | - |  |
|  |  |  | Water | City water | Proportion exposed = 49 | 35-63 | - |  |
|  |  |  | Water | Drank untreated/raw water | Proportion exposed = 11 | 4-24 | - |  |
|  |  |  | Water | Private well | Proportion exposed = 31 | 19-46 | - |  |
|  |  |  | Water | Swam in/gone into ocean, lake, river, pool, hot tub | Proportion exposed = 46 | 31-61 | - |  |
|  |  |  | Water | Used an in-home treatment system for drinking water | Proportion exposed = 24 | 13-38 | - |  |
| **Case-control** | **^[2]^NicLochlainn, 2018**  Reports the 3-year overall study results only | **Netherlands** | Food | Ate tomatoes | aOR=0.6 | 0.5-0.8 | 0.001 | MVA |
|  |  |  | Food | BBQ food | aOR=1.8 | 1.4-2.3 | 0.001 |  |
|  |  |  | Person to person | Household person-to-person transmission | aOR=2.2 | 1.7-3.0 | 0.001 |  |
| **Case-control** | **De Gooyer, 2017** | **Australia** | Person to person | Household member with diarrhoea | aOR=12.6 | 2.13-75.1 | 0.006 | MVA |
|  |  |  | Water | Drank bottled water | aOR=6.31 | 1.39-28.7 | 0.017 | MVA |
|  |  |  | Water | Waterpark | aOR=36.9 | 3.12-435 | 0.004 | MVA |
|  |  |  | Water | Spa use | aOR=26.4 | 1.47-472 | 0.026 | MVA |
|  |  |  | Water | Recreational water | OR=3.18 | 1.15-8.79 | 0.023 | UVA |
|  |  |  | Water | Waterpark | OR=73.5 | 6.74-802 | 0.0001 | UVA |
|  |  |  | Water | Public pool | OR=1.07 | 0.44-2.60 | 0.89 | UVA |
|  |  |  | Water | Private pool | OR=1.96 | 0.65-5.96 | 0.225 | UVA |
|  |  |  | Water | Natural bodies of water | OR=1 | 0.31-3.25 | 1 | UVA |
| **Case-control** | **Fournet, 2013 (1) (NL study)** | **Netherlands** | Water | Drank bottled mineral water | aOR=2.72 | 1.10-6.76 | 0.03 | MVA |
|  |  |  | Animals | Contact with farm animals | Difference in Proportions (case v control) = 33% vs 40% | - | 0.3 | UVA |
|  |  |  | Travel | Travel | Difference in Proportions (case v control) = 36% vs 22% | - | 0.03 | UVA |
|  |  |  | Water | Drank tap water on daily basis | Difference in Proportions (case v control) = 67% vs 78% | - | 0.07 | UVA |
|  |  |  | Water | Exposure to swimming pool, sea, river or lake | Difference in Proportions (case v control) = 70% vs 66% | - | 0.56 | UVA |
| **Case-control** | **Valderrama, 2009** | **USA** | Food | Consumption of produce from farm/farm stand | aOR=0.2 | 0.1-0.9 | - | MVA |
|  |  |  | Person to person | Attending social event | aOR=0.4 | 0.1-0.9 | - |  |
|  |  |  | Person to person | Contact with child in child-care or in diapers | aOR=3.8 | 1.5-9.6 | - |  |
|  |  |  | Water | Drinking untreated water from lake, river, or stream | aOR=8.0 | 1.3-48.1 | - |  |
|  |  |  | Water | Exposure to any recreational water | aOR=4.6 | 1.4-14.6 | - |  |
| **Case-case** | **Fournet, 2013 (2) (UK study)** | **UK** | Animals | Dog ownership | Difference in Proportions (year 2012 vs 2009-11) = 46% vs 25% | - | 0.02 | UVA |
|  |  |  | Food | Ate food prepared outside the home | Difference in Proportions (year 2012 vs 2009-11) = 32% vs 4% | - | 0.001 |  |
|  |  |  | Travel | Travel | Difference in Proportions (year 2012 vs 2009-11) = 54% vs not reported | - | 0 |  |
|  |  |  | Water | Bottled water | Difference in Proportions (year 2012 vs 2009-11) = 11% vs 10% | - | 0.44 |  |
|  |  |  | Water | Swimming pool use | Difference in Proportions (year 2012 vs 2009-11) = 18% vs 37% | - | 0.02 |  |
| **Case-case** | **Pintar, 2009** | **Canada** | Animals | Visited a farm | OR=1.6 | 1-2.5 | 0.032 | UVA |
|  |  |  | Case characteristics | Age 0-5 | OR=2.8 | 0.84-9.1 | 0 | UVA |
|  |  |  | Case characteristics | Age 6-12 | OR=5.5 | 1.7-18 | 0 | UVA |
|  |  |  | Case characteristics | Age 13-17 | -- | 0-0 | 0 | UVA |
|  |  |  | Case characteristics | Age 18-24 | OR=1.7 | 0.38-7.9 | 0 | UVA |
|  |  |  | Case characteristics | Age 25-39 | OR=2.2 | 0.65-7.4 | 0 | UVA |
|  |  |  | Case characteristics | *Age 40-59* | *Ref grp* | *--* | *--* | *--* |
|  |  |  | Case characteristics | Age >60 | OR=0.8 | 0.15-4.6 | 0 | UVA |
|  |  |  | Food | Ate a ready-to-eat product | OR=2.1 | 0.5-9.3 | 0.276 | UVA |
|  |  |  | Food | Ate at a fast food restaurant | OR=1.5 | 0.7-3.4 | 0.273 | UVA |
|  |  |  | Food | Ate at a food vendor | OR=2.8 | 0.6-13 | 0.186 | UVA |
|  |  |  | Food | Killed own food | OR=2.3 | 0.5-10 | 0.245 | UVA |
|  |  |  | Food | Meat from butchers | OR=0.8 | 0.2-3.5 | 0.516 | UVA |
|  |  |  | Outdoor activities/environmental exposure | Hiking, camping, or canoeing | OR=2.1 | 0.8-5.7 | 0.135 | UVA |
|  |  |  | Outdoor activities/environmental exposure | Lived on a farm | OR=2.5 | 1.1-5.6 | 0.023 | UVA |
|  |  |  | Person-to-Person | Attending social event | OR=0.23 | 0.05-0.99 | 0 | MVA |
|  |  |  | Person-to-Person | Family member with diarrhoea | OR=2.86 | 1.28-6.38 | 0 | MVA |
|  |  |  | Person-to-Person | Non-family member with diarrhoea | OR=2.1 | 0.64-6.87 | 0 | MVA |
|  |  |  | Person-to-Person | Attended a social gathering | OR=0.3 | 0.1-1.1 | 0.068 | UVA |
|  |  |  | Person-to-Person | Family member ill | OR=3 | 1.4-6.3 | 0.003 | UVA |
|  |  |  | Person-to-Person | Non-family member with diarrhoea | OR=1.9 | 0.7-5.3 | 0.184 | UVA |
|  |  |  | Season | Autumn vs spring as ref grp | OR=5.8 | 0.74-46 | 0 | UVA |
|  |  |  | Season | Summer vs spring as ref grp | OR=5.6 | 0.74-42 | 0 | UVA |
|  |  |  | Season | Winter vs spring as ref grp | OR=1.4 | 0.08-22 | 0 | UVA |
|  |  |  | Water | Municipal water supply | OR=2.43 | 1.05-5.65 | 0.04 | MVA |
|  |  |  | Water | Swimming in natural water | OR=2.91 | 1.14-7.38 | 0 | MVA |
|  |  |  | Water | Municipal water supply (vs private) | OR=2.2 | 1-4.8 | 0 | UVA |
|  |  |  | Water | Swimming in untreated water lake or river | OR=3.1 | 1.4-6.7 | 0.004 | UVA |
|  |  |  | Water | Swimming pool | OR=1.9 | 0.8-4.8 | 0.154 | UVA |
|  |  |  | Water | Went swimming | OR=3.8 | 1.8-8.2 | 0.001 | UVA |
| **Serological** | **Becker, 2015** | **USA** | Case characteristics | Age (year increments from 6-49 years) | aOR=1.06 | 1.05-1.07 | <0.001 | MVA |
|  |  |  | Case characteristics | Black ethnicity (vs White) | aOR=1.88 | 1.42-2.53 | 0.001 |  |
|  |  |  | Case characteristics | Country of birth Mexico (vs USA) | aOR=2.96 | 1.99-4.25 | 0.001 |  |
|  |  |  | Case characteristics | Country of birth Other (vs USA) | aOR=2.27 | 1.53-3.28 | 0.001 |  |
|  |  |  | Case characteristics | Hispanic ethnicity (vs White) | aOR=1.76 | 1.38-2.28 | 0.001 |  |
|  |  |  | Case characteristics | Other ethnicity (vs White) | aOR=2.13 | 1.1-4.07 | 0.04 |  |
|  |  |  | Other | Annual income more than $45,000 (vs <$25,000) | aOR=0.61 | 0.41-0.9 | 0.03 |  |
|  |  |  | Other | Food adequacy (not enough vs enough) | aOR=1.31 | 0.67-2.59 | 0.65 |  |
|  |  |  | Other | Poverty income ratio high (vs low) | aOR=0.75 | 0.51-1.1 | 0.14 |  |
|  |  |  | Water | Untreated drinking water (vs treated) | aOR=1.19 | 0.97-1.45 | 0.13 |  |
| **Serological** | **^[3]^Tollestrup, 2014 (a)** | **USA** | Animals | Handled livestock | OR=0.53 | 0.29-0.97 | -- | MVA |
|  |  |  | Water | Private wastewater system/well | OR=1.98 | 1.11-3.55 | -- |  |
|  | **Tollestrup, 2014 (b)** | **USA** | Water | Private wastewater system/well | OR=1.28 | 0.74-2.21 | -- | MVA |
|  |  |  | Water | Plumbing work done in home | OR=2.11 | 1.10–4.03 | -- |  |
|  |  |  | Animals | Handled pets | OR=2.83 | 1.24–6.49 | -- |  |
|  |  |  | Case characteristics | Age 18-39 | *Ref grp* | *--* | -- |  |
|  |  |  | Case characteristics | Age 40–49 | OR=0.89 | 0.40–2.01 | -- |  |
|  |  |  | Case characteristics | Age 50–59 | OR=1.8 | 0.80–4.03 | -- |  |
|  |  |  | Case characteristics | Age 60+ | OR=4.2 | 1.79–9.89 | -- |  |
|  |  |  | Case characteristics | College graduate (vs not) | OR=0.37 | 0.19–0.72 | -- |  |
|  | **Tollestrup, 2014 (c)** | **USA** | Water | Private wastewater system/well | OR=0.85 | 0.49–1.46 | -- | MVA |
|  |  |  | Case characteristics | Age 18-39 | *Ref grp* | *--* | -- |  |
|  |  |  | Case characteristics | Age 40–49 | OR=1.76 | 0.74–4.18 |  |  |
|  |  |  | Case characteristics | Age 50–59 | OR=1.92 | 0.83–4.43 |  |  |
|  |  |  | Case characteristics | Age 60+ | OR=3.69 | 1.61–8.46 |  |  |
