## Supplementary figures and images for "Exposures associated with sporadic *Cryptosporidium* infection in industrialised countries: a systematic review"

### PRISMA Flowchart

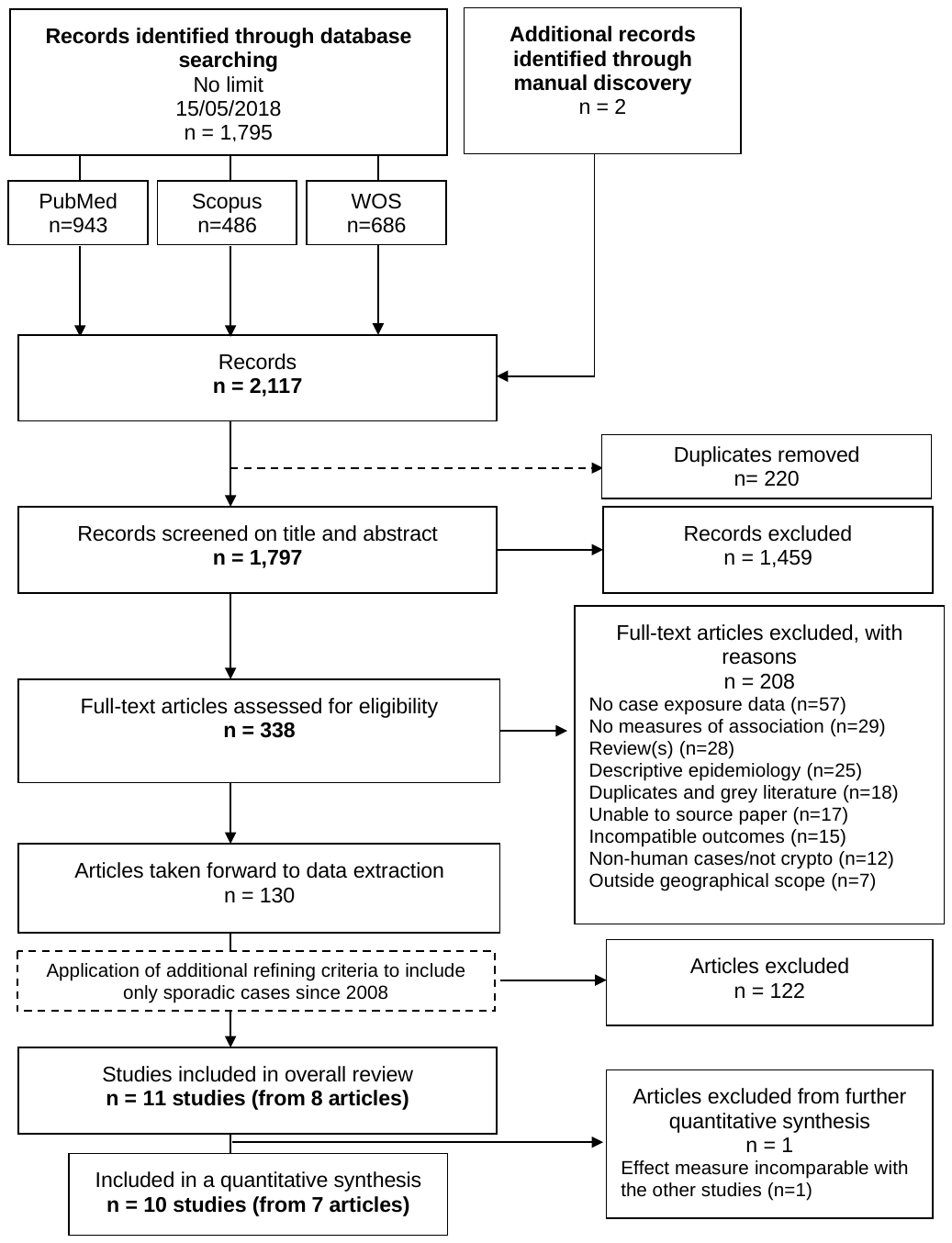

*Figure 1: PRISMA diagram showing manuscript capture and inclusion*
